# Heart Block in the Patients with COVID-19

**DOI:** 10.1101/2022.01.05.22268779

**Authors:** Md Ripon Ahammed, Medha Sharath, Mehul Sinha, Cristina Sestacovschi, Varadha Retnakumar, Naisargee Solanki

## Abstract

**Background:** Since the emergence of the SARS COV-19 pandemic, multiple extrapulmonary manifestations of the virus have been reported from around the world. Cardiovascular complications including arrhythmias in patients with COVID-19 have been described in multiple studies. Our aim was to review various case reports detailing the new onset of heart block in COVID-19 patients and to summarise the clinical course of these patients.

**Methods:** We systematically reviewed all reports published and indexed in PubMed, Scopus, and Embase between March 2020 to May 2021, analyzing the relation between the demographics of the patients, pre-existing comorbidities, and the progression of heart block in patients infected with COVID-19.

**Results:** We identified and included in this study 30 relevant articles describing 49 COVID-19 patients with heart block. Among them, 69.3% (n=34) of patients suffered from at least one comorbidity. 36.73% (n=18) of the patients showed spontaneous resolution of the heart block. Conversely, 63.26% (n=31) of the patients had persistent heart block, out of which 16.33% (n=8) and 42.86% (n=21) were implanted with a temporary and permanent pacemaker respectively. The reported mortality rate was 22.45% (n=11) during hospitalization. We noted that 45.45% (n=5) of the patients who died had complete heart block. 24.49% (n=12) of the patients in the studies we reviewed were suspected of having myocarditis. However, none were confirmed with MRI or cardiac biopsy.

**Conclusions:** Additional research is necessary to unearth the mechanism of development of heart block in COVID-19 patients as well as its implications on the clinical course and prognosis. Physicians must be aware of the importance of monitoring patients hospitalized for COVID-19 for arrhythmias including heart blocks, especially in the presence of comorbidities. Early detection can improve the prognosis of the patient.

## Background

Since the first reports of atypical pneumonia in Wuhan province of China, the coronavirus disease 2019 (COVID-19) has evolved into a major world health issue. Although pneumonia is the most significant manifestation of severe acute respiratory syndrome coronavirus-2 (SARS-CoV-2) infection^1^, a range of extra-pulmonary manifestations have been reported, such as venous thromboembolism, cardiac complications, gastrointestinal abnormalities, acute kidney insult, hepatocellular injury, hyperglycemic state, neurological illnesses and dermatological complications^2^. 14.1% of hospitalized COVID-19 cases have been reported to develop cardiovascular abnormalities which mainly include acute coronary syndrome, arrhythmia, and acute heart failure^3^; although patients with fulminant myocarditis, pericarditis, cardiac tamponade, Takotsubo cardiomyopathy, and cor pulmonale have also been described^4–7^. It is recognized that cardiac involvement in COVID-19 leads to a worse prognosis^8^. Ventricular arrhythmias, atrial arrhythmias, sinus bradycardia, heart block, ST-segment changes, and prolonged QTc have all been reported in COVID-19 patients. Meta-analyses have estimated the arrhythmia incidence rate in patients hospitalized for COVID-19 to be from 6.9% to 10.3% ^9,10^.

Heart block is the least documented form of arrhythmia that is seen in COVID-19 patients^10^ and the incidence, nature, and impact of this type of arrhythmia among COVID-19 patients is yet to be studied. Hence, the aim of this systematic review is to understand potential patterns of heart block, association with pre-existing comorbidities, and the effect on the clinical course including the outcome of heart block in this group of patients.

## METHODS

### Literature Search

2 researchers (RA, and MS) independently conducted the search and selection of the studies. We included case reports and case series published from March 2020 to May 2021. The systematic search was carried out in May 2021 utilizing PubMed (Medline), Scopus, and Embase. The keywords that we searched for include: heart block OR arrhythmia OR dysrhythmia OR bradyarrhythmia OR brady-arrhythmia OR atrioventricular block OR atrioventricular block AND novel coronavirus OR new coronavirus OR nCoV OR Cov-2 OR Cov2 OR COVID AND case OR case report OR case reports OR case series.

### Screening, Inclusion and Exclusion Criteria

There was no pre-specified exposure/ intervention, comparison groups, or outcomes as we intended to summarize all available case reports of COVID-19 and heart block. Titles, abstracts, and full-texts were screened by RA and MS independently. Letters to the editor, commentaries, systematic reviews, meta-analyses, and duplicates were excluded. Figure 1 is the PRISMA flow diagram which shows the selection process of our study. Our search criteria yielded the identification of 437 articles. 407 articles were excluded after screening. 2 articles were further excluded after review since they did not describe heart block or COVID-19, and 1 article was excluded since it was a systematic review. 3 articles were added after a manual search. Finally, 30 eligible articles were selected for our systematic review.

**Fig 1:**
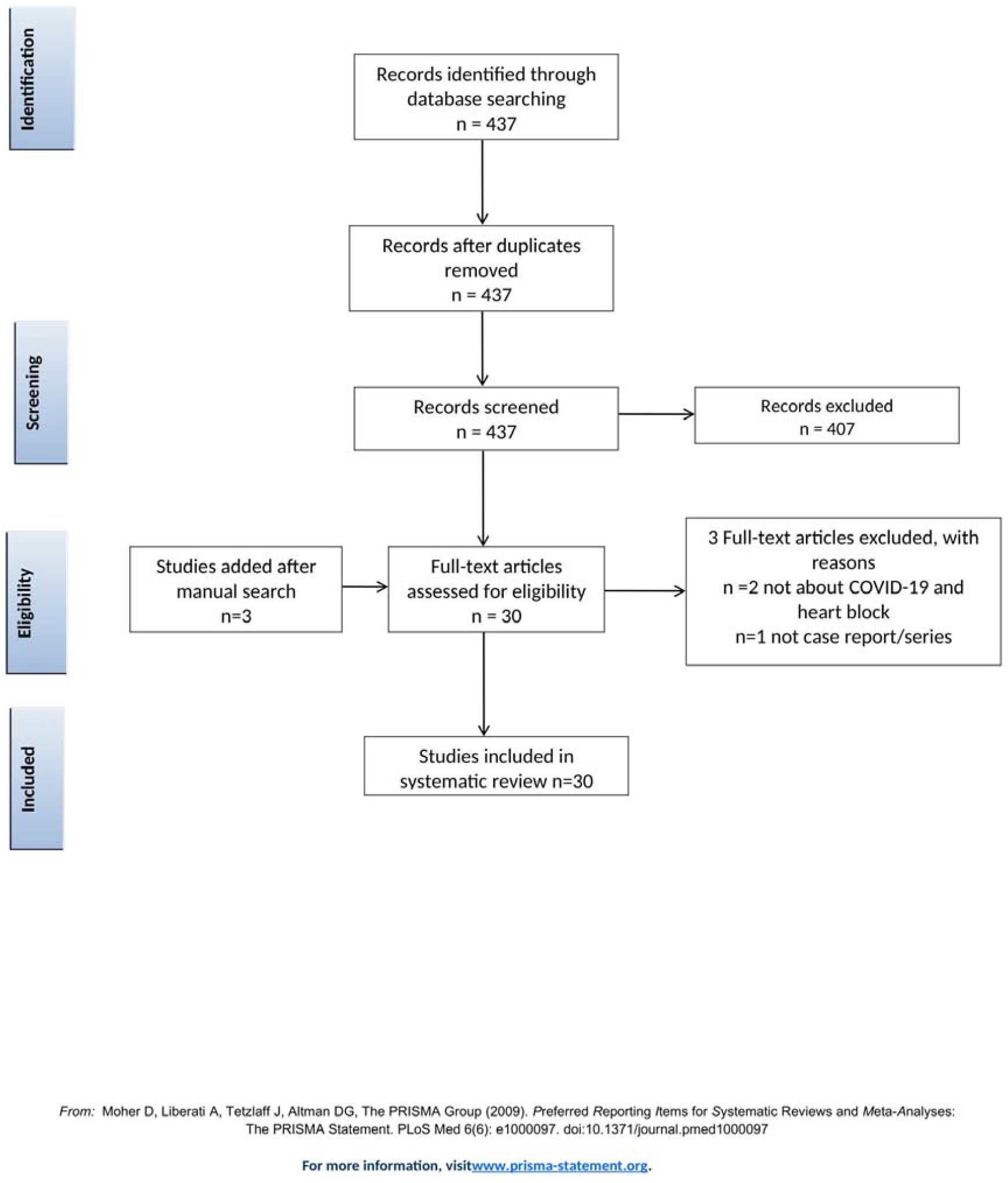
Prisma Flow Chart

### Extraction of Data

Data from included studies were extracted by reviewers and verified by using a standard form. Data regarding patient demographics; comorbidities; medications; baseline ECG abnormalities; laboratory findings; onset, duration, characteristics, and resolution of heart block; as well as details of pacemaker implant were collected in an excel sheet. The data was verified twice to check for any inaccuracies or discrepancies.

## RESULTS

We identified and included in this study 30 relevant articles describing 49 patients. Our selected studies include 7 case series and 23 case reports. Most patients described were from the USA, followed by India and Iran. Tables 1, 2, and, 3 detail the data extracted from our selected studies.

**TABLE 1:**
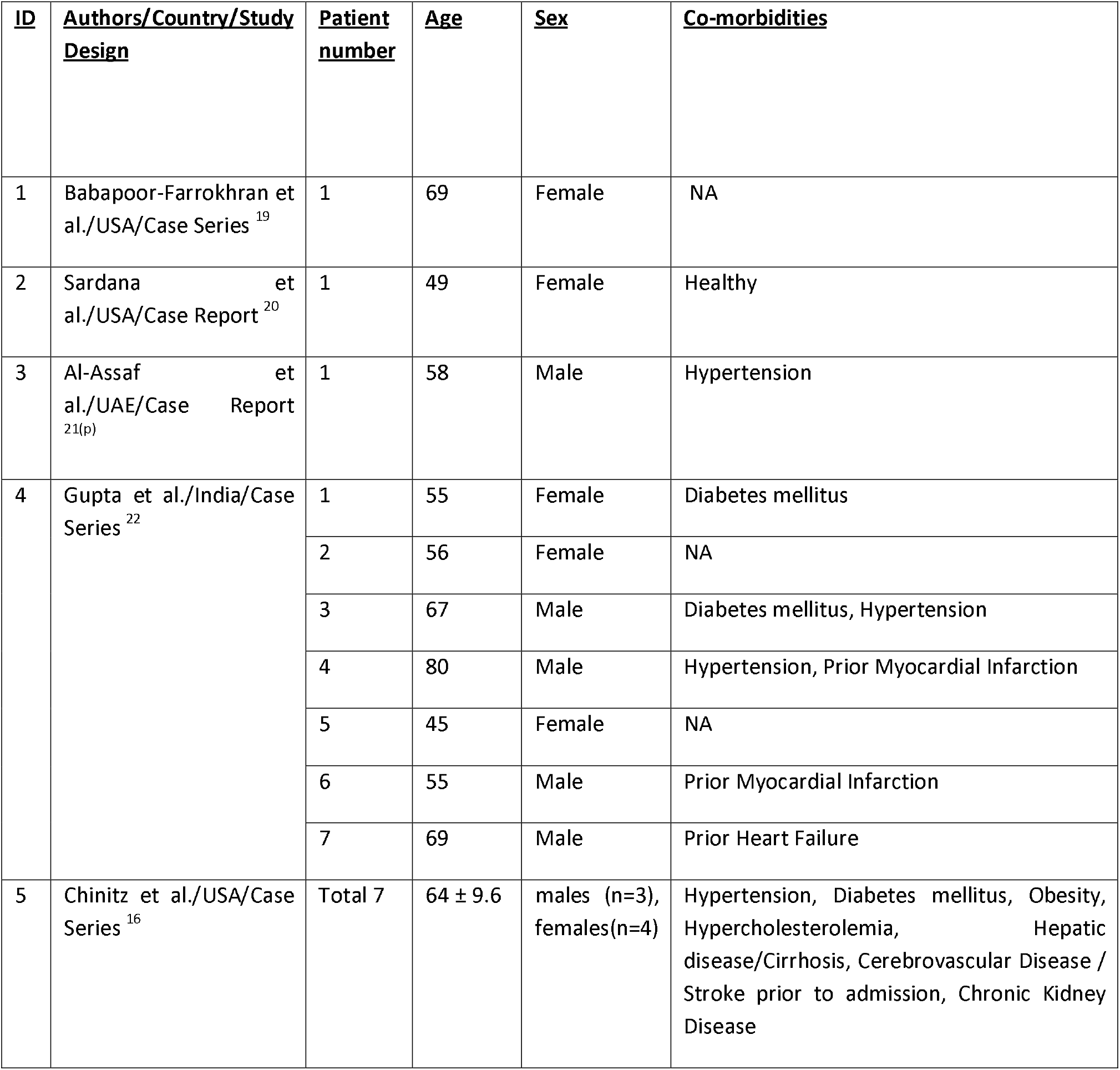
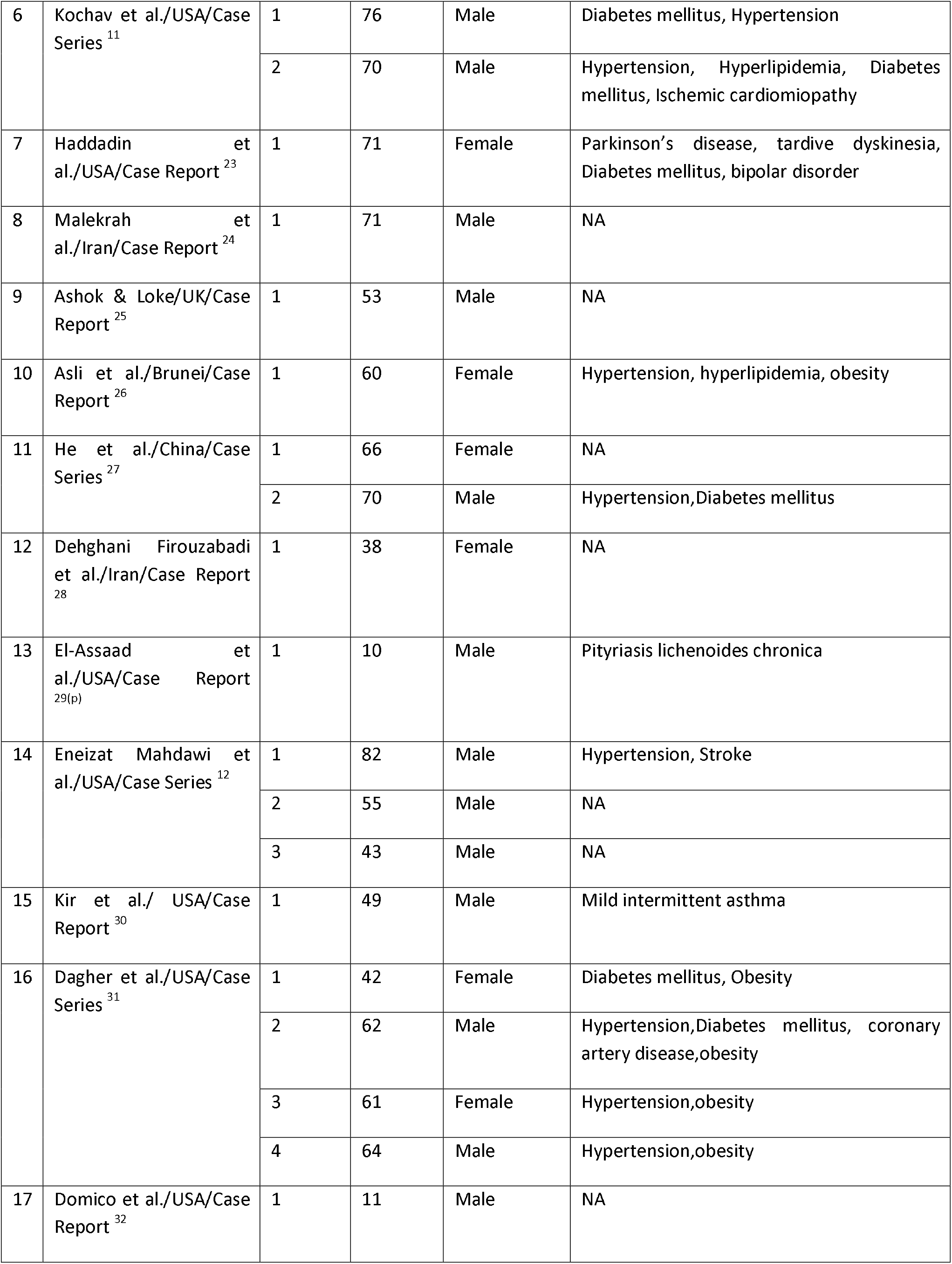
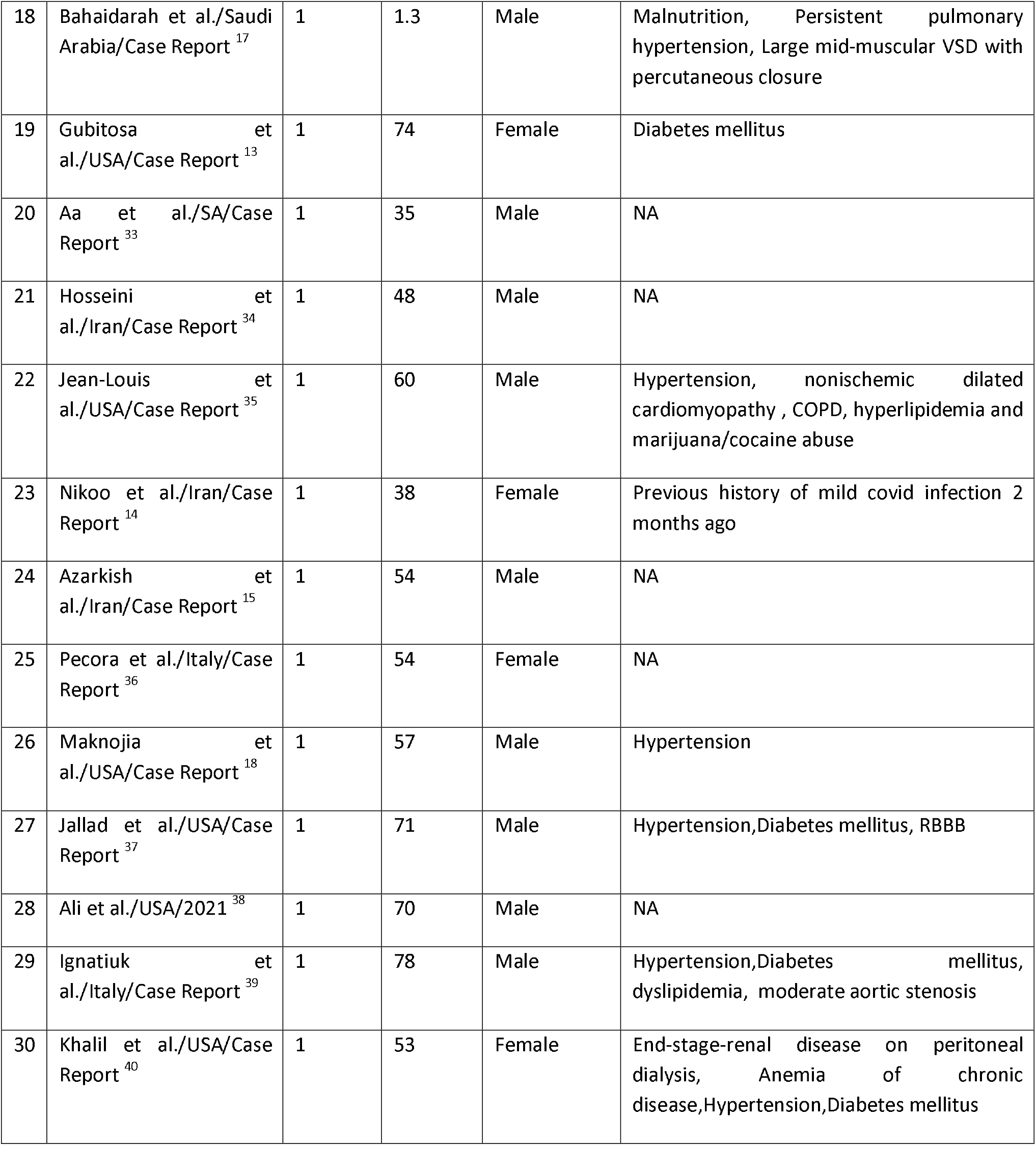
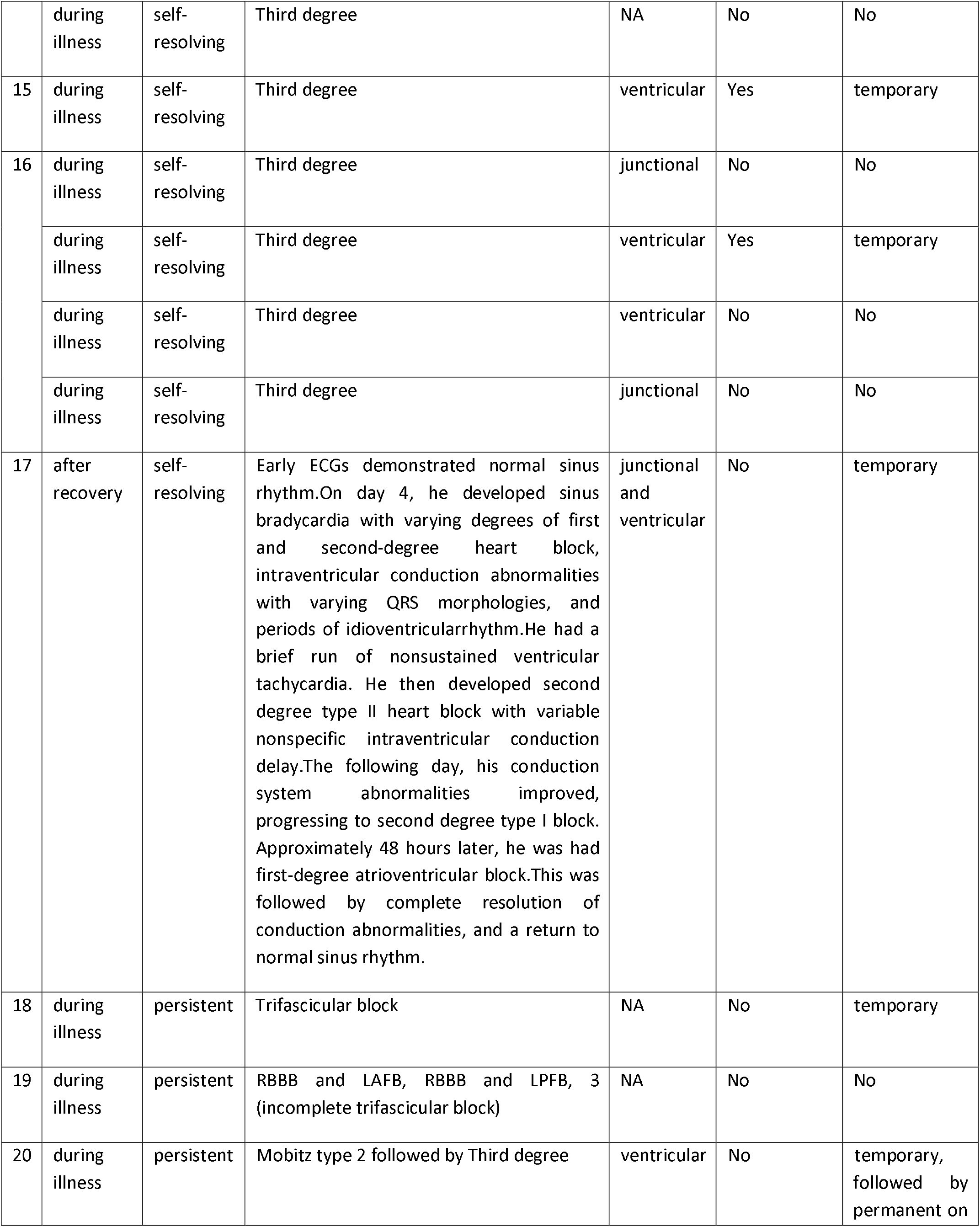
Characteristics of included studies

**TABLE 2:**
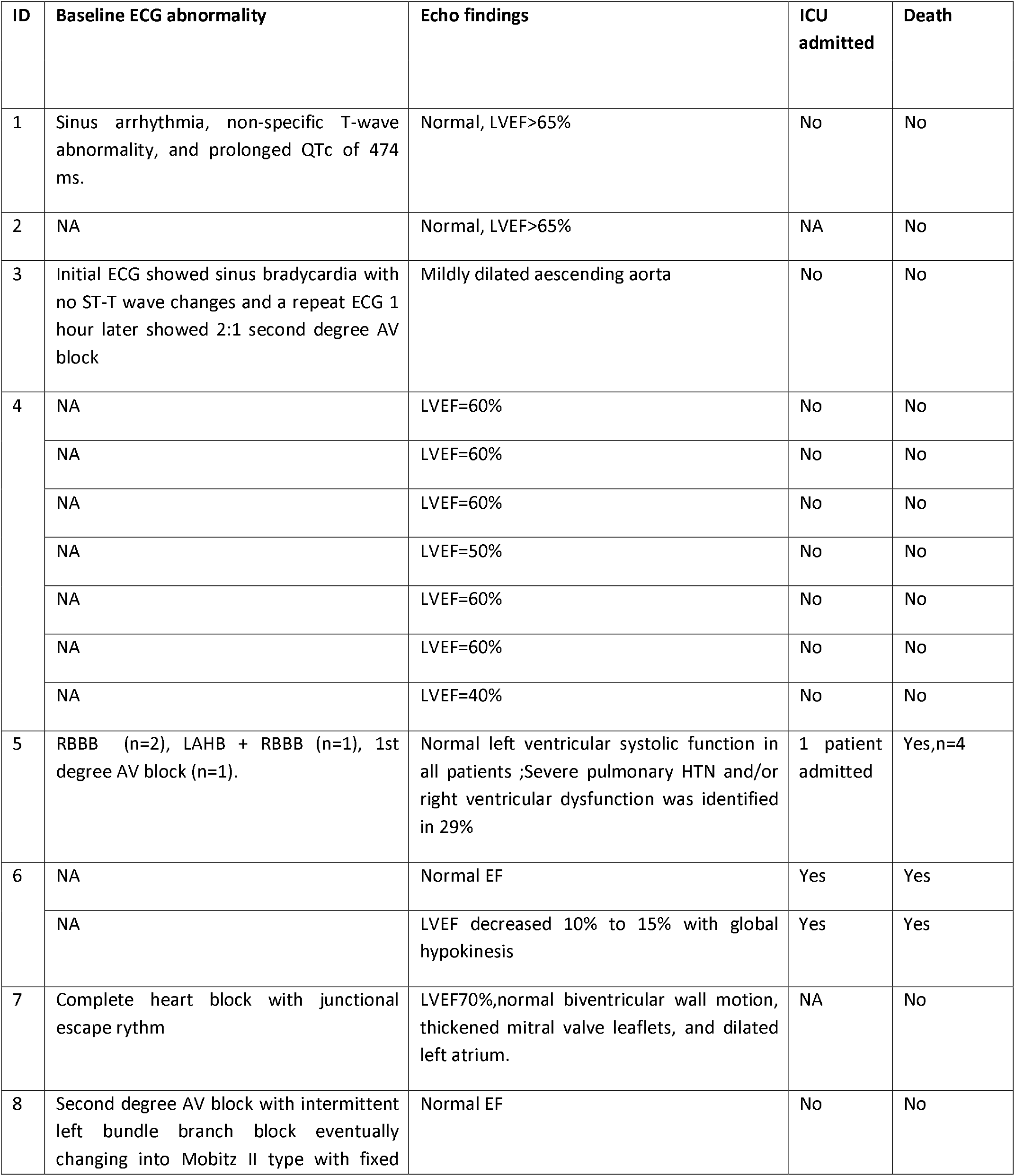
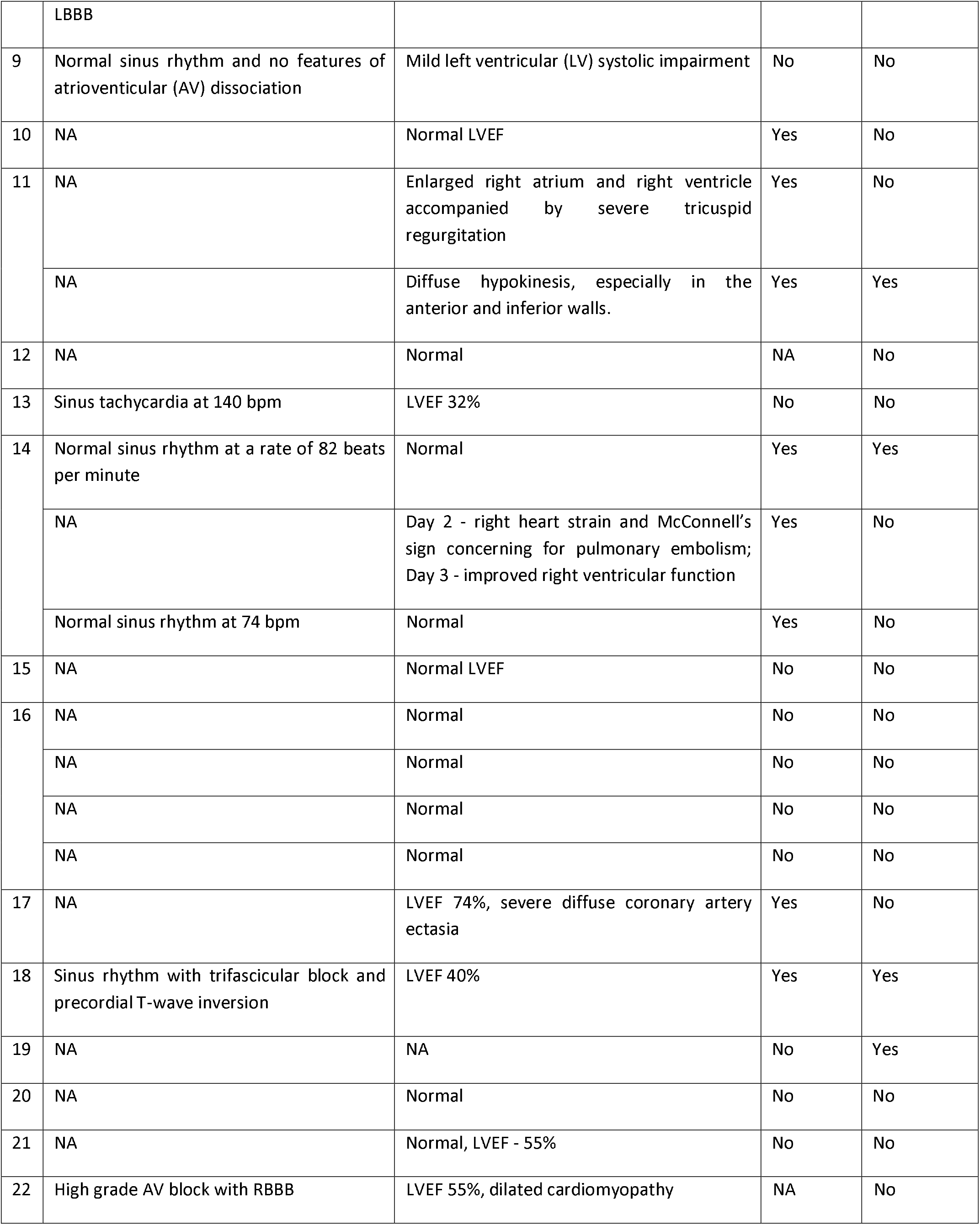
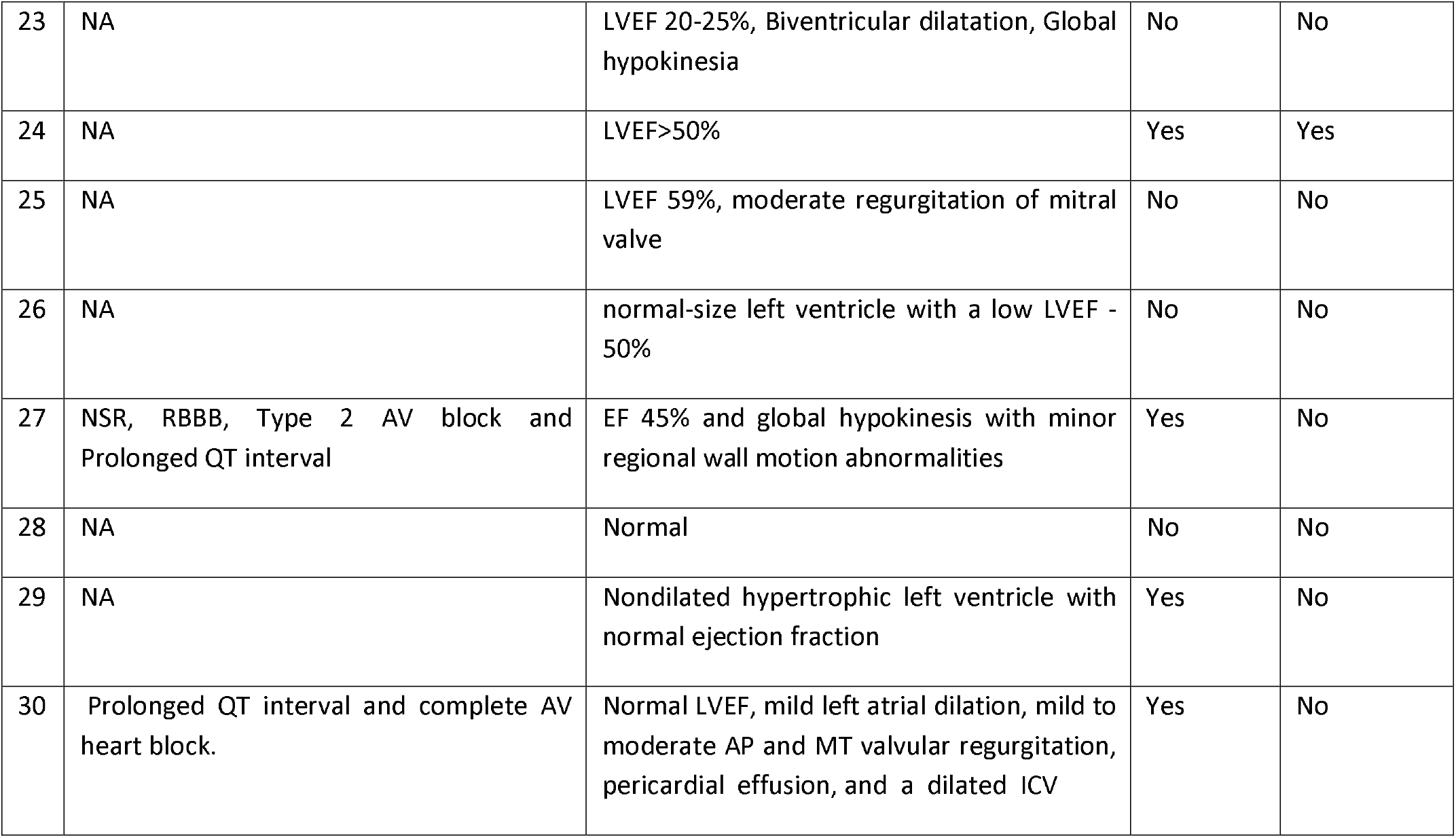
Outcome of the included patients

**TABLE 3:**
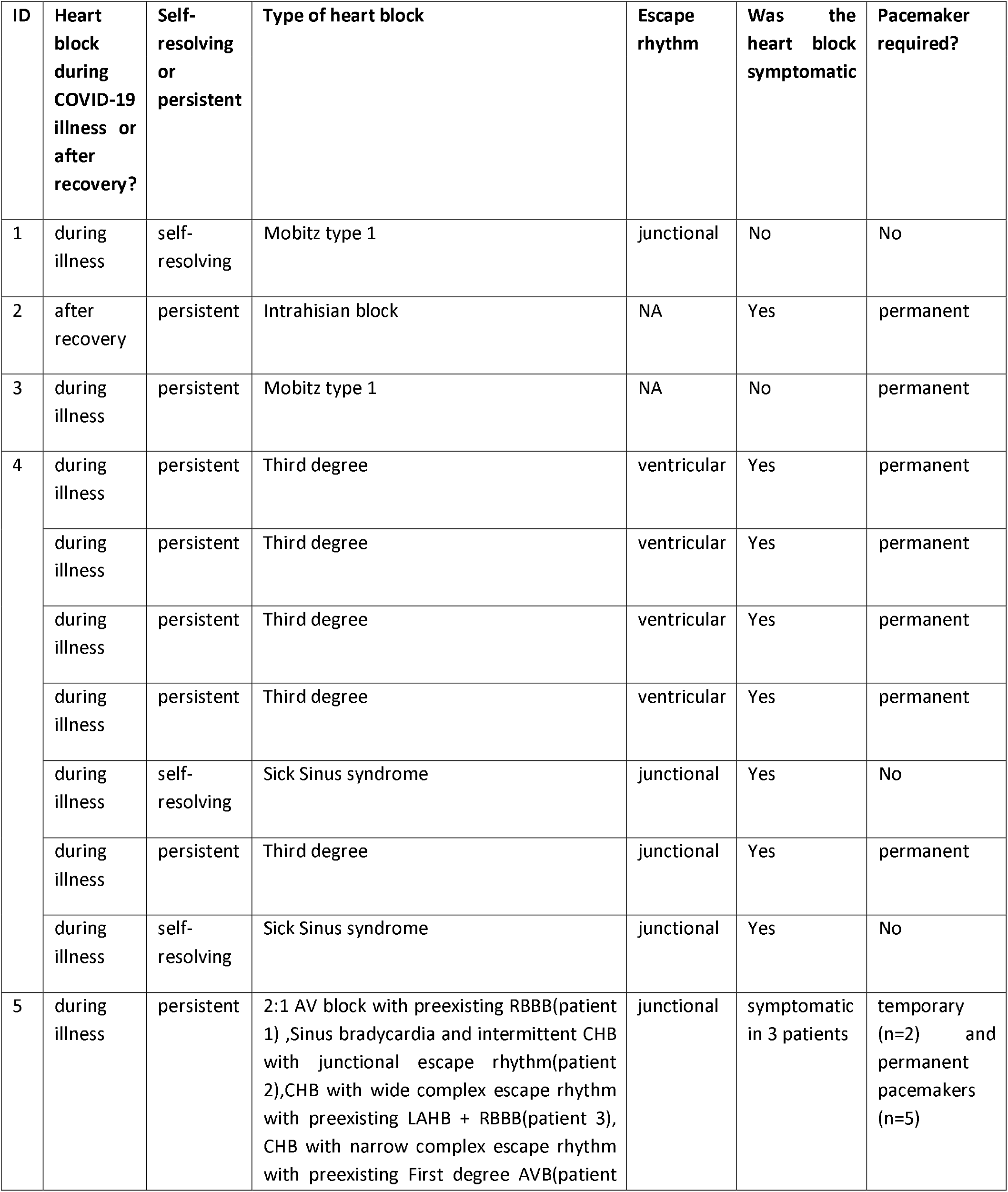
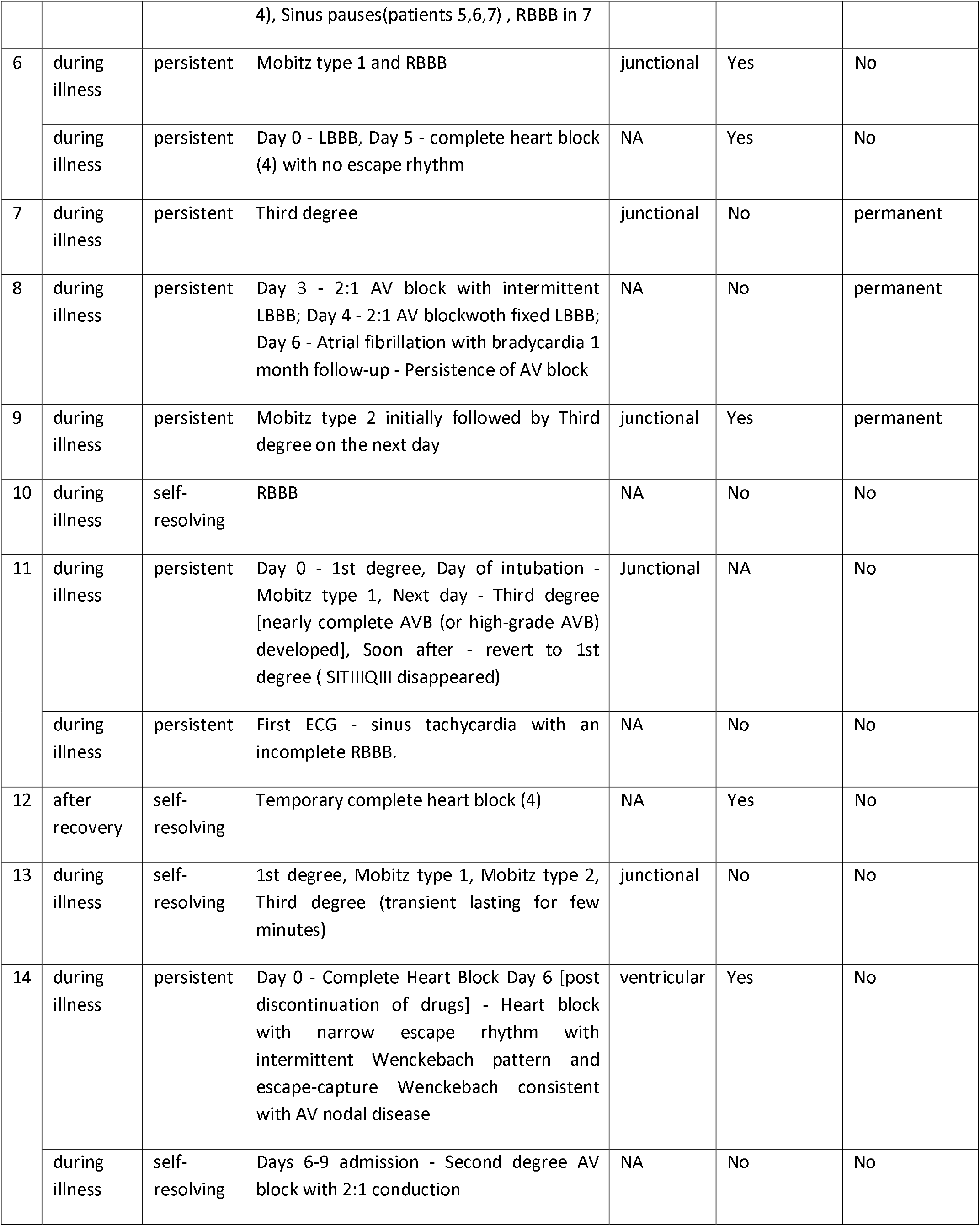
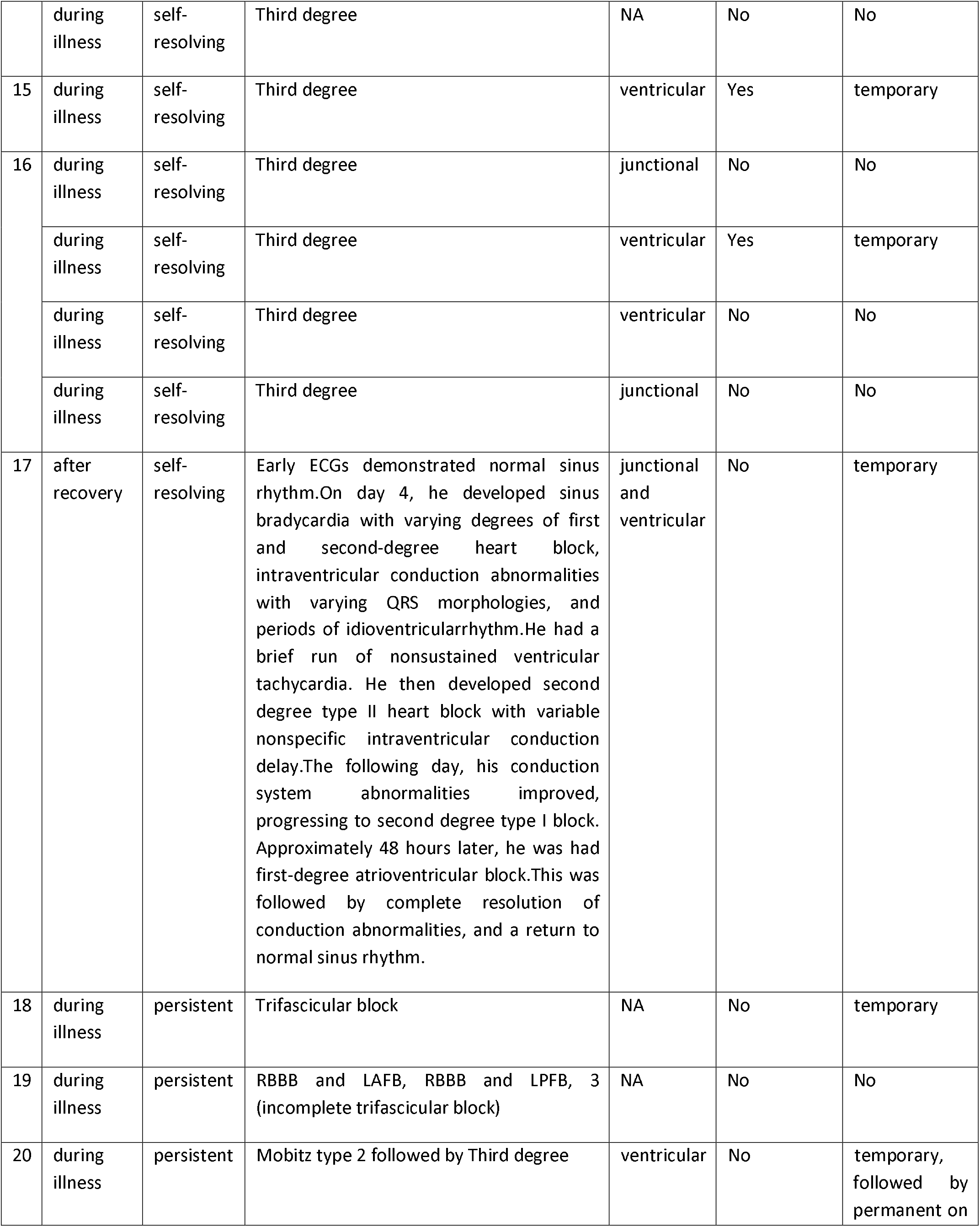
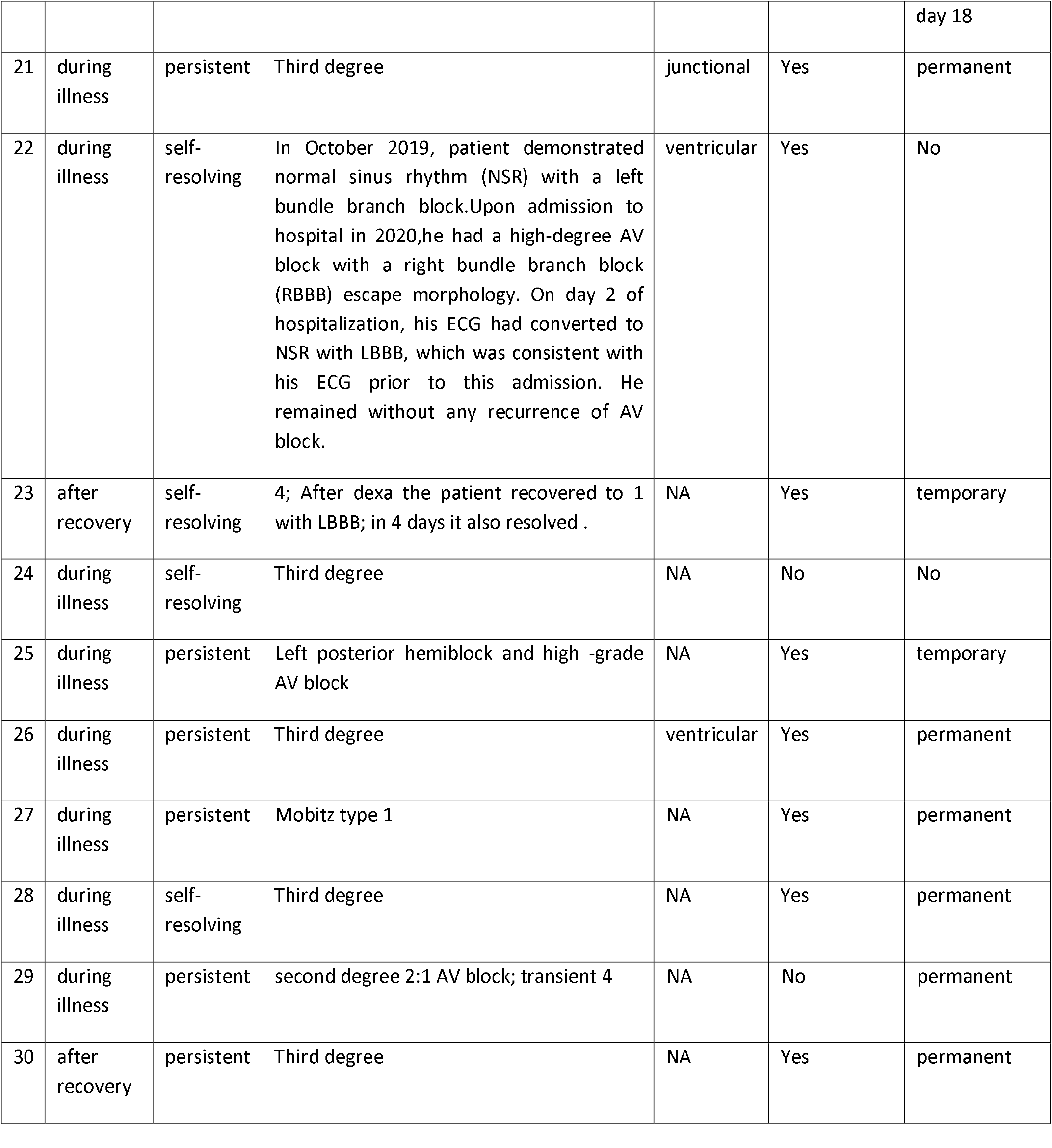
Different types of heart block in the studied patients

Overall the age of the patients varied from 16 months to 82 years (Mode=64) and were predominantly males (61.2%; n=30). In 69.3% (n=34) of the cases, patients suffered from at least one comorbidity: 64.7% (n=22) of them had arterial hypertension (men being affected twice as much as women), 52.9% (n=18) had Diabetes mellitus, 17.6% (n=6) had hyperlipidemia, 11.7% (n=4) reported a previous myocardial infarction, and 8.8% (n=3) had a previous stroke. The duration of active COVID-19 infection varied from 3 to 90 days, with most cases lasting around 14 days. 97.9% (n=48) of all patients were hospitalized but only around 1/3 of them were admitted into ICU (30.6%). 32.65% (n=16) of the hospitalized patients required concurrent mechanical intubation/ventilation.

Regarding laboratory findings, 20.4% (n=10) of the total number of patients diagnosed with heart block had reduced left ventricular ejection fraction (LVEF) in the echocardiogram. Among these, LVEF was mildly reduced (LVEF=54-40%) in 50% (n=5) and severely reduced (LVEF<35%) in 10% (n=1). CRP was elevated in 55.4% (n=27) of the patients, and 37% (n=10) of them tested positive for HsCRP. WBC count was raised in 20.4% (n=10) of the patients. Myocardial involvement was measured in terms of increased Troponin I, which was raised in 22.4% (n=11) of the cases.

Our review found that 89.8% (n=44) of the patients described in the included studies developed heart block during hospitalization for COVID-19 infection, while 10.20% (n=5) of the patients developed heart block following recovery. The type of heart block varied throughout the studies we reviewed, but the most common was third-degree heart block present in 61.22% (n=30) of the patients. Junctional escape rhythm was seen in 30.61% (n=15), and Ventricular escape rhythm was seen in 26.53% (n=13) of the patients. The heart block was symptomatic in 53.06% (n=26), the most common symptom being dyspnea. 36.73% (n=18) of the patients showed spontaneous resolution of the heart block. On the other hand, 63.26% (n=31) of the patients had persistent heart block, out of which 16.33% (n=8) and 42.86% (n=21) were implanted with a temporary and permanent pacemaker, respectively. 24.49% (n=12) of the patients in the studies we reviewed were suspected of having myocarditis. However, none were confirmed with MRI or cardiac biopsy. The reported mortality rate in our study was 22.45% (n=11) during hospitalization. We noted that 45.45% (n=5) of the patients who died had complete heart block^11–17^. The other types of heart block reported in the other patients who died include Mobitz type 1 heart block with RBBB^11^, sinus pauses and sinus arrest, sinus tachycardia with an incomplete right bundle branch block (RBBB)^18^, trifascicular block^17^, RBBB and left anterior fascicular block (LAFB), RBBB, left posterior fascicular block (LPFB) and incomplete trifascicular block^13^.

### Changes in the type of heart block during the clinical course

We have found that 61.22% of the patients in our case studies developed a third-degree heart block. Some other ECG findings reported after COVID-19 infection include first-degree heart block (n=4); Mobitz type 1 (n=7); Mobitz type 2 (n=10); atrial fibrillation (AF) with bradycardia (n=1); Intrahisian block (n=1) and Trifasicular block (n=1). Six patients experienced RBBB and four patients experienced LBBB. Sick sinus syndrome was also detected in 2 patients ^22^. Many patients had changes in the type of heart block during the course of illness. Alabdulgader et al. described a patient who had persistent heart block and received a temporary pacemaker; this was later replaced with a permanent pacemaker on day 18 of hospitalization ^33^. Pecora et al described a patient with left posterior hemiblock and high-grade AV block, which resolved with implantation of a temporary pacemaker ^36^. 2 patients developed Mobitz type 2 heart block, which was followed by 3rd-degree complete heart block ^13,41^. Azakirsh et al illustrated a patient who developed Mobitz type 2 AV block (2:1) with intermittent LBBB on the 3rd day of hospitalization; this evolved on the fourth day to Mobitz type 2 AV block (2:1) with fixed LBBB, and on the sixth day to AF with bradycardia. The persistence of AV block was observed one month after discharge ^15^. Domico et al reported a child with a high-grade heart block, which resolved with temporary pacing. Upon admission, the patient’s ECGs exhibited normal sinus rhythm. On the fourth day, the patient developed sinus bradycardia along with varying degrees of first and second-degree heart block, episodes of idioventricular rhythm, and intraventricular conduction abnormalities associated with fluctuating morphologies in QRS complexes. Consequently, the patient experienced a transient nonsustained ventricular tachycardia (VT) followed by a second-degree type II heart block with inconsistent nonspecific intraventricular conduction delay. On day 5, the irregularities of the conduction system improved and settled into a second-degree type I block. On day 7, he demonstrated a first-degree AV block which was followed by complete resolution of conduction abnormalities and return to sinus rhythm ^32^.

One patient showed inferior and precordial leads ST-segment elevation on day 35 of hospitalization. In addition, there was a progressive increase in ST-elevation amplitude which transformed into a triangular QRS-ST-T waveform. Two attacks of multifocal ventricular tachycardia were observed during the development of ST-segment elevation ^27^.

Additional ECG findings reported in the included studies are sinus bradycardia ^19^, sinus tachycardia^26^, sinus pauses in 3 patients ^16^, and QT prolongation ^24,32,37,40^.

## DISCUSSION

Every day brings about a new discovery about the pathogenesis and clinical manifestations of COVID-19. According to the fast-growing availability of scientific literature on this topic, the heart appears to be one of the elective targets of the virus. It is known that SARS-CoV enters into human cells via Angiotensin-Converting Enzyme 2 (ACE-2) receptors which are found in alveolar epithelial cells and endothelium of arteries and veins. Multiple hypotheses have been made to explain the mechanism of heart blocks in COVID-19 infection. In autopsy studies, SARS-CoV-2 virus and inflammatory infiltrate have been found in the myocardium, which implies direct viral invasion of the heart. ACE-2 downregulation decreases the action of angiotensin 1–7 leading to increased synthesis of inflammatory mediators like TNFα, CRP, and TGFβ which produces a cytokine storm. TGFβ induces interstitial fibrosis, which damages cardiac architecture. Troponin elevation and contractility dysfunction occur in the setting of severe hypoxia due to inflammatory damage or hypercoagulability. These phenomena initiated by the virus can occur in parallel with direct viral damage and interact with each other, enhancing their effect ^8,42^.

In our study, only 24.49% of the patients were suspected of myocarditis. However, none were confirmed with MRI or cardiac biopsy. Kir et al. reported one patient with no evidence of myocardial involvement indicated by normal levels of cardiac enzymes and no findings on ECG, who developed a self-resolving 3rd-degree heart block. It is postulated that COVID-19 triggered subclinical myocarditis may have given rise to high-degree AV block in this patient ^30^. The mechanism of heart block in other cases is poorly understood. In our review, all the patients who died after developing heart block were on mechanical ventilation in the intensive care unit.

Deterioration of any pre-existing diseases in the conduction system such as AV node disease, bundle branch blocks, or His-Purkinje system disorder can cause new advanced blocks, leading to poor clinical outcomes ^34^. Another big role in COVID-19 progression is played by comorbidities and risk factors. Cardiovascular diseases, hypertension, diabetes mellitus, renal disease, liver disease, cerebrovascular disease, obesity, hyperlipidemia, and smoking history have a crucial impact on disease progression and complications ^8^. In our review, hypertension and diabetes mellitus were present in the majority of patients who passed away due to heart block during COVID-19 infection. Since many of these risk factors are modifiable, lifestyle changes, early diagnosis, and management of comorbidities must be considered for better outcomes in COVID-19 patients.

Pathophysiological, histological, and imaging data ^8,43^ indicate that SARS-CoV-2 could induce tissue damage, which would predispose patients to recurrent cardiac issues long-term after discharge. However, the long-term impact of COVID-19 induced heart blocks on late cardiac manifestations are not well studied, thus leading to poor clinical guidance regarding remote cardiac follow-up after discharge.

### Limitations

One of the limitations of our systematic review was the restricted number of studies and size of the sample population. We have only included studies published until May 2021. Future reports of heart block in COVID-19 patients might reveal new relations between the severity of the infection and the prognosis of patients who develop heart block.

The resolution of ECGs changes after discontinuation of HCQ in a case suggests a causal relationship between hydroxychloroquine and ECG abnormalities. Thus the role of certain medications, like hydroxychloroquine, on the development of arrhythmias in the presence of COVID-19 infection must also be further investigated ^26^.

Also, the presence of COVID-19 induced myocarditis was not confirmed via biopsy or MRI in any of the patients who developed heart block in our included studies.

### Conclusion

The exact pathogenesis of heart block in COVID-19 patients must be further explored. More research is required to determine whether myocarditis due to COVID-19 is associated with the development of heart blocks. It is also needed to be studied further whether heart block is an independent risk factor for a worse prognosis in COVID-19 cases.

Patients hospitalized for COVID-19 must be continuously monitored for arrhythmias including heart blocks, especially in the presence of comorbidities. Physicians must be made aware of these cardiac manifestations of the infection, as early detection can shorten the hospital stay and improve the prognosis of the patient.

## Data Availability

All data produced in the present study are available upon reasonable request to the authors

## Contributions

MRA contributed to research idea, design of the study, collecting data, statistical analysis and interpretation, writing the first draft, writing and editing the final manuscript. MSh contributed to study design, data extraction, statistical analysis and interpretation, writing the first draft. MS, CS, VR and NS contributed to data extraction, statistical analysis & interpretation and writing the first manuscript.

## Acknowledgment

To Larkin Community Hospital, FL, USA for supporting and providing our research platform.

## Ethical Approval

Not applicable

